# Pan-cancer proteogenomic landscape of whole-genome doubling reveals putative therapeutic targets in various cancer types

**DOI:** 10.1101/2024.04.16.24305805

**Authors:** Eunhyong Chang, Hee Sang Hwang, Kyu Jin Song, Kwoneel Kim, Min-Sik Kim, Se Jin Jang, Kwang Pyo Kim, Sungyong You, Joon-Yong An

**Author notes:** **Correspondence:** (Joon-Yong An).

## Abstract

**Background:** Whole-genome doubling (WGD) is prevalent in cancer and drives tumor development and chromosomal instability. Driver mutations in mitotic cell cycle genes and cell cycle upregulation have been reported as the major molecular underpinnings of WGD tumors. However, the underlying genomic signatures and regulatory networks involved in gene transcription and kinase phosphorylation remain unclear. Here, we aimed to comprehensively decipher the molecular landscape underlying WGD tumors.

**Methods:** We performed a pan-cancer proteogenomic analysis and compared 10 cancer types by integrating genomic, transcriptomic, proteomic, and phosphoproteomic datasets from the Clinical Proteomic Tumor Analysis Consortium (CPTAC). We also integrated the cancer dependency data of each cancer cell line and the survival properties of each cancer patient to propose promising therapeutic targets for patients with WGD.

**Results:** Our study delineated distinct copy number signatures characterizing WGD-positive tumors into three major groups: highly unstable genome, focal instability, and tetraploidy. Furthermore, the analysis revealed the heterogeneous mechanisms underlying WGD across cancer types with specific structural variation patterns. Upregulation of the cell cycle and downregulation of the immune response were found to be specific to certain WGD tumor types. Transcription factors (TFs) and kinases exhibit cancer-specific activities, emphasizing the need for tailored therapeutic approaches.

**Conclusion:** This study introduces an integrative approach to identify potential TF targets for drug development, highlighting BPTF as a promising candidate for the treatment of head and neck squamous cell carcinoma. Additionally, drug repurposing strategies have been proposed, suggesting potential drugs for the treatment of WGD-associated cancers. Our findings offer insights into the heterogeneity of WGD and have implications for precision medicine approaches for cancer treatment.

## Background

Whole-genome doubling (WGD) is prevalent across cancer subtypes, promoting tumor development and generating chromosomal instability (CIN). WGD plays an important role in tumorigenesis by cushioning the deleterious mutations and rapidly accumulating genetic abnormalities. However, WGD is also associated with tolerance to genomic instability, which leads to cell death ^1–3^. Previous studies have investigated the genomic alterations associated with WGD, revealing that *TP53* mutations and defects in the E2F-mediated G1 arrest are common in WGD-positive tumors ^1^. Moreover, gene expression studies have highlighted the enrichment of genes involved in cellular proliferation, mitotic spindle formation, and DNA repair, whereas inflammatory pathways are downregulated in WGD-positive tumors ^2^. These findings depict the overall pan-cancer characteristics of WGD; however, the role of WGD can be highly heterogeneous ^2,4–7^.

Recent proteomic studies have delineated novel mechanisms underlying diverse cancer subtypes ^8^. These investigations have revealed associations between certain multiomic subtypes that are indicative of WGD. For example, in head and neck squamous cell carcinoma (HNSCC), a distinct molecular subtype has been characterized by high CIN and upregulated cell cycle pathways at both the proteome and phosphoproteome levels ^9^. Similar WGD-associated subtypes have been detected in non-small cell lung cancer (NSCLC), endometrial cancer, breast cancer (BRCA), colon cancer, and glioblastoma (GBM) ^10–14^. Despite mounting evidence implicating WGD-related subtypes across multiple cancers, a pan-cancer multiomics investigation focusing on WGD remains elusive. In addition, the proteomic features and kinase activities governing WGD in cancer are yet to be elucidated. Furthermore, the development of therapeutic strategies that specifically target WGD-positive tumors remains an unmet need.

In this study, we aimed to conduct a pan-cancer proteogenomic analysis to delineate the genomic and proteomic landscapes of WGD across ten types of cancer by integrating genomic, transcriptomic, proteomic, and phosphoproteomic data sets. Our objective was to characterize the molecular pathways, transcription factor (TF) regulation, and kinase phosphorylation networks enriched in association with the WGD. Finally, we explored the potential drug targets and repositioning strategies for patients with WGDs.

## Results

### Pan-cancer analysis to identify CN signatures underlying WGD

We sought to explore the proteogenomic features associated with WGD by analyzing comprehensive genomic, transcriptomic, proteomic, and phosphoproteomic datasets obtained from the Clinical Proteomic Tumor Analysis Consortium (CPTAC) (**Figure 1A**). The dataset comprised 1,060 patients representing 10 types of cancer: breast cancer (BRCA) ^12^, clear cell renal cell carcinoma (CCRCC) ^15^, colon adenocarcinoma (COAD) ^13^, glioblastoma (GBM) ^14^, high-grade serous carcinoma (HGSC) ^16,17^, head and neck squamous cell carcinoma (HNSCC) ^9^, lung adenocarcinoma (LUAD) ^11^, lung squamous cell carcinoma (LSCC) ^10^, pancreatic ductal adenocarcinoma (PDAC) ^18^, and uterine corpus endometrial carcinoma (UCEC) ^12^. By determining the WGD status of each sample, we observed a bimodal distribution of patients, indicating the existence of two distinct groups of cancers irrespective of the cancer type (**Figure 1B**). Consistent with previous estimates, approximately 42% of the tumors (440 of 1,060 samples) exhibited at least 1 occurrence of WGD during their evolutionary process^1,2^. We also identified substantial variability in the occurrence of WGD across different tumor types, with HGSC showing the highest prevalence (83%, 65/78 samples) and PDAC showing the lowest (9.4%, 13/139 samples) (**Figure 1C**).

**Figure 1.**
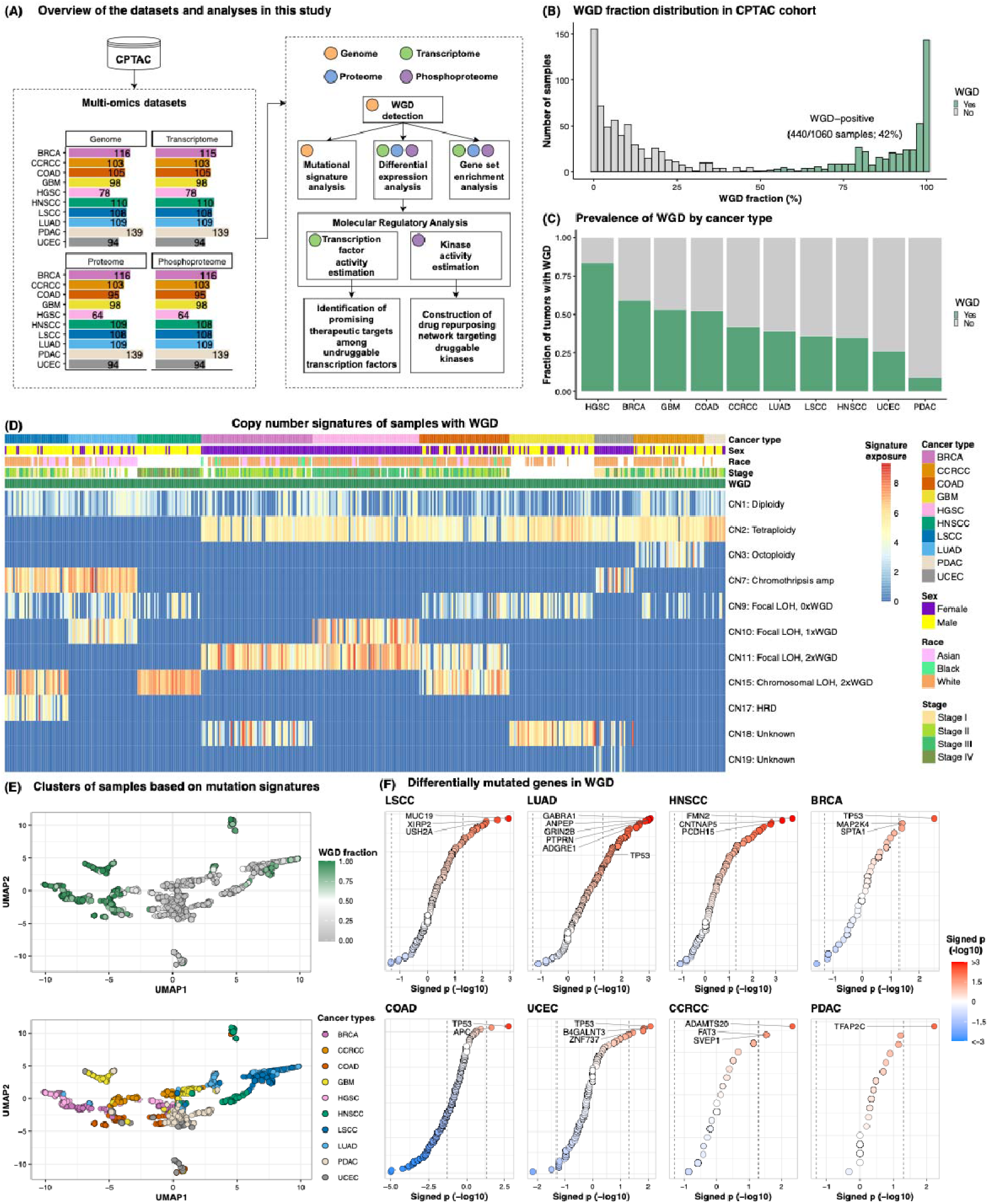
Copy number signatures associated with WGD. **(A)** Schematic representation of the multi-omics datasets collected for each cancer type and the subsequent analysis pipeline. BRCA, breast cancer; CCRCC, Clear cell renal cell carcinoma; COAD, colon adenocarcinoma; GBM, Glioblastoma; HGSC, High-grade serous carcinoma; HNSCC, Head and neck squamous cell carcinoma; LSCC, Lung squamous cell carcinoma; LUAD, Lung adenocarcinoma; PDAC, Pancreatic ductal adenocarcinoma; UCEC, Uterine corpus endometrial carcinoma. **(B)** Distribution of the WGD fraction within the CPTAC cohort, displaying a bimodal pattern. **(C)** Prevalence of WGD by cancer type. **(D)** Heatmap based on copy number signature exposure values in samples with WGD. Copy number signatures were derived from the Catalogue Of Somatic Mutations In Cancer (COSMIC). **(E)** UMAP plot based on signature exposure values of single-base-substitution, double-base-substitution, indel, and copy number alteration. Each dot indicates samples and is colored based on WGD fraction (top) and cancer types (bottom). **(F)** Dot plot depicting differentially mutated genes in WGD in each cancer type. The position of each dot along the x-axis and the color of the dots indicate signed p-values obtained from Fisher’s exact test. Only the top three significant genes were labeled in each cancer type, except for COAD and PDAC, for which only two and one significant genes, respectively, were noted. In LUAD, *PTPRN, GRIN2B, ANPEP*, and *ADGRE1* genes exhibited the same p-values, and *TP53* was additionally labeled.

Mutation signature analyses demonstrated that WGD was associated with specific copy number signatures (**Table S1B**). Based on the 25 CN signature values of the WGD-positive samples, different cancer types exhibited varying combinations of CN signatures, suggesting that the underlying mechanisms of WGD might be distinct across cancer types (**Figure 1D**). In patients with LSCC, those with WGD showed significant enrichment for CN7 (FDR = 3.11×10^−5^), indicating chromothripsis amplification, as well as for CN15 (FDR = 2.87×10^−2^), signifying chromosomal loss-of-heterozygosity (LOH) with twice-genome-doubling (**Figure S1A; Table S1B**). Similarly, patients with HNSCC showed significant enrichment of CN15 in WGD-positive samples (FDR = 4.63×10^−6^). Patients with LUAD showed enrichment of the CN7 signature (FDR = 5.39×10^−4^). WGD in BRCA and HGSC correlated significantly with CN11, a signature of focal LOH, with two WGD events (BRCA, FDR=1.71×10^−5^; HGSC, FDR=2.68×10^−13^), suggesting that WGD in these malignancies occurred within a focally unstable genomic context. A majority of WGD samples from CCRCC, COAD, GBM, PDAC, and UCEC were found to be significantly enriched for CN2 (FDR < 0.05), indicating tetraploidy. Based on these observations, we defined three distinct WGD status in LSCC, LUAD, and HNSCC as “WGD type 1,” that in BRCA and HGSC as “WGD type 2,” and that in CCRCC, COAD, GBM, PDAC, and UCEC as “WGD type 3.” The WGD status was present in a coherent grouping of samples, wherein cancer types with similar CN signatures exhibited spatial proximity (**Figure 1E**).

We further evaluated the driver mutations associated with WGD in each tumor type. Despite previous reports suggesting an enrichment of *TP53* mutations in WGD tumors across diverse cancers ^1,2^, we identified a significant enrichment of *TP53* mutations exclusively in the WGD-positive samples of BRCA, COAD, UCEC, and LUAD (p < 0.05; Fisher’s exact test) (**Figure 1F; Table S1C**). Although no common gene mutations were identified, we observed that over 100 mutations were significantly enriched in association with WGD in LSCC, LUAD, and HNSCC (WGD type 1), whereas other cancer types exhibited fewer than 15 significant gene mutations associated with WGD. As a highly unstable genome has been linked to a higher tumor mutational burden (TMB) ^19,20^, we speculated that the TMB would be higher in WGD type 1. We found significantly higher TMB in WGD-positive samples than in WGD-negative samples across 10 cancer types (p = 3.12×10^−4^; Wilcoxon rank-sum test) (**Figure S1A**), and in LSCC, LUAD, HNSCC, and BRCA when compared across individual cancer types (p < 0.05; Wilcoxon rank-sum test) (**Figure S1B**). WGD-positive samples in COAD exhibited significantly lower TMB than WGD-negative samples (p = 2.13×10^−4^; Wilcoxon rank-sum test). Nevertheless, *TP53* and *APC* mutations remained significantly enriched in samples with WGD in COAD (*TP53*, p = 1.75×10^−3^; *APC*, p = 2.31×10^−2^; Fisher’s exact test) (**Figure 1F**). This is consistent with the findings of prior investigations suggesting an association between *APC* mutations and aneuploidy in COAD ^21–23^. These observations imply that genomic instability is a catalyst for WGD in LSCC, LUAD, and HNSCC, whereas *TP53* and *APC* mutations may serve as primary drivers of WGD in COAD. In summary, our findings revealed distinct CN signatures of WGD, allowing us to define three types of WGD.

### Enrichment of distinct pathways in WGD in each cancer type

Previous studies have reported activation of the cell cycle pathway and inactivation of the immune response pathway in tumors with WGD ^1,2,24,25^. Given the various CN signatures underlying WGD across cancer types, we conducted a sample-level pathway enrichment test with 1,060 samples and examined the biological pathways enriched in WGD (**Figure 2A**). Overall, WGD-positive tumors were significantly affected by several pathways (**Table S2A**), which were subsequently categorized into four major pathway groups: cell motility, immune response, cell cycle, and metabolism. WGD type 1 tumors, characterized by a highly unstable genome, showed significant upregulation of the cell cycle and downregulation of immune response pathways (**Figure 2B and 2C**), which is consistent with previous reports ^1,2,24,25^. In contrast, WGD type 2 tumors showed significant enrichment in the dTTP metabolism pathway, which is responsible for DNA synthesis and maintenance ^26,27^ (p < 0.01; Wilcoxon rank-sum test), and the DNA endoreduplication pathway, a known mechanism inducing WGD ^28,29^ (p < 0.05; Wilcoxon rank-sum test) (**Figure S2A and S2B**). Among other cancer types, WGD tumors in COAD showed significant activation of the Wnt signaling pathway, possibly attributed to *APC* mutation, in line with the findings of previous studies ^30–32^ (p = 2.09×10^−3^; Wilcoxon rank-sum test) (**Figure S2C**). These results emphasize that upregulation of the cell cycle and downregulation of the immune response are specific to WGD type 1 tumors, suggesting diverse functional attributes of WGD across distinct cancer types.

**Figure 2.**
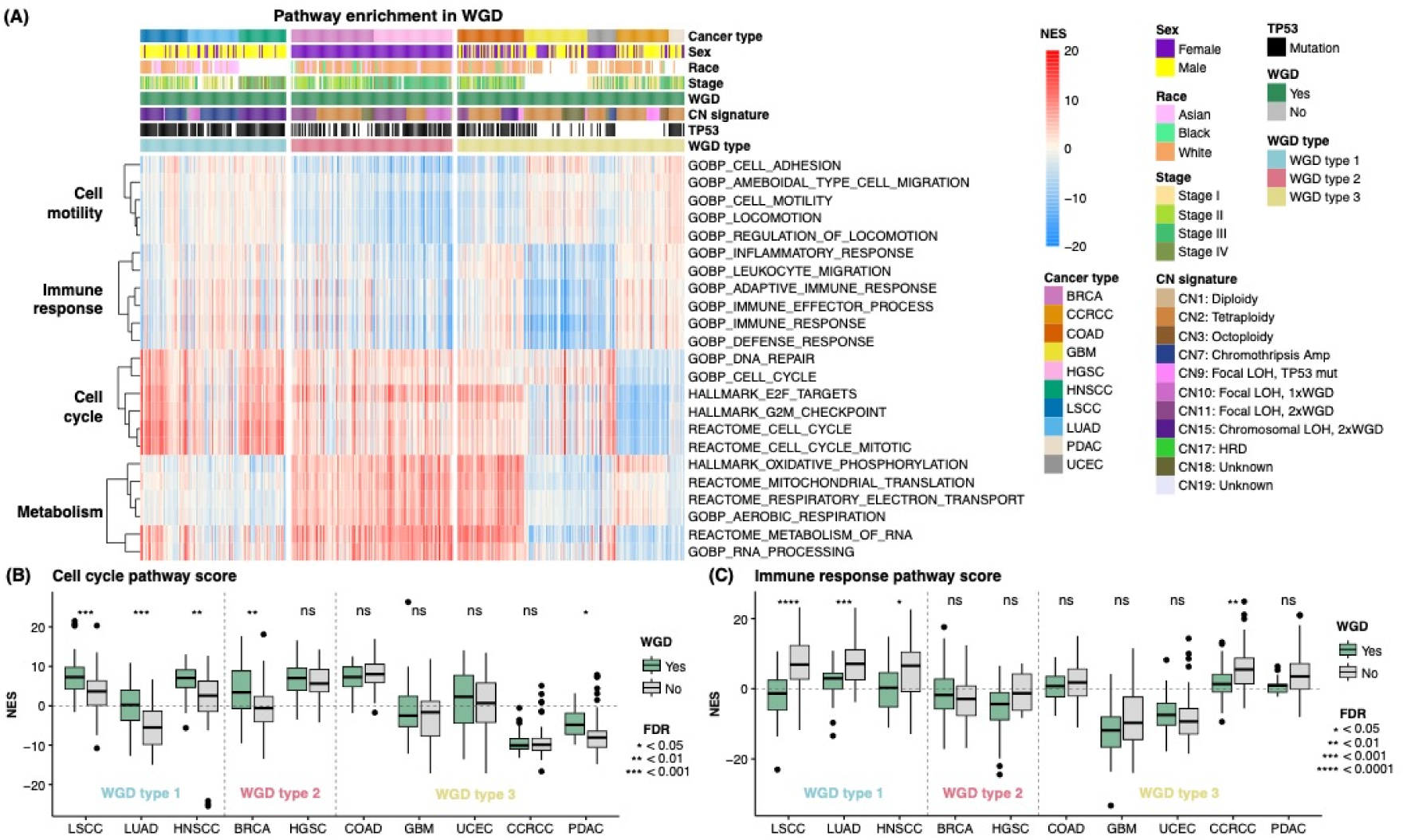
Pathway enrichment in WGD-positive tumors. **(A)** Normalized enrichment scores (NES) of pathways related to cell motility, immune response, cell cycle, and metabolism in WGD-positive tumors. Pathways exhibiting significance (FDR < 0.05) and a log fold-change greater than 0.25 are shown. **(B–C)** Boxplot comparing NES score of cell cycle pathway (HALLMARK_E2F_TARGETS) and immune response pathway (GOBP_IMMUNE_RESPONSE) between WGD-positive and WGD-negative tumors in individual cancer types.

### WGD-specific TFs as potential therapeutic targets

To identify potential therapeutic targets for treating WGD across various tumor types, we first estimated TF activity using the TF-target gene interaction network database ^33^ and the gene expression levels of target genes (see Methods). Out of the 1,134 TFs analyzed, the E2F family and MYC TFs showed significant activation (FDR < 0.05) in pan-cancer WGD tumors, indicating the role of cell cycle regulation in WGD pathophysiology ^34,35^ (**Figure 3A; Table S3A**). In contrast, the TFs that were significantly downregulated in the WGD-positive tumors were predominantly associated with immune response pathways. While different TFs were activated in WGD-positive tumors across different cancer types (**Figure 3B**), E2F1, E2F2, E2F3, E2F4, and MYC were common TFs with significant activation in five cancer types (LSCC, LUAD, HNSCC, BRCA, and PDAC) that showed upregulation of the cell cycle pathway (**Figure 2B**).

**Figure 3.**
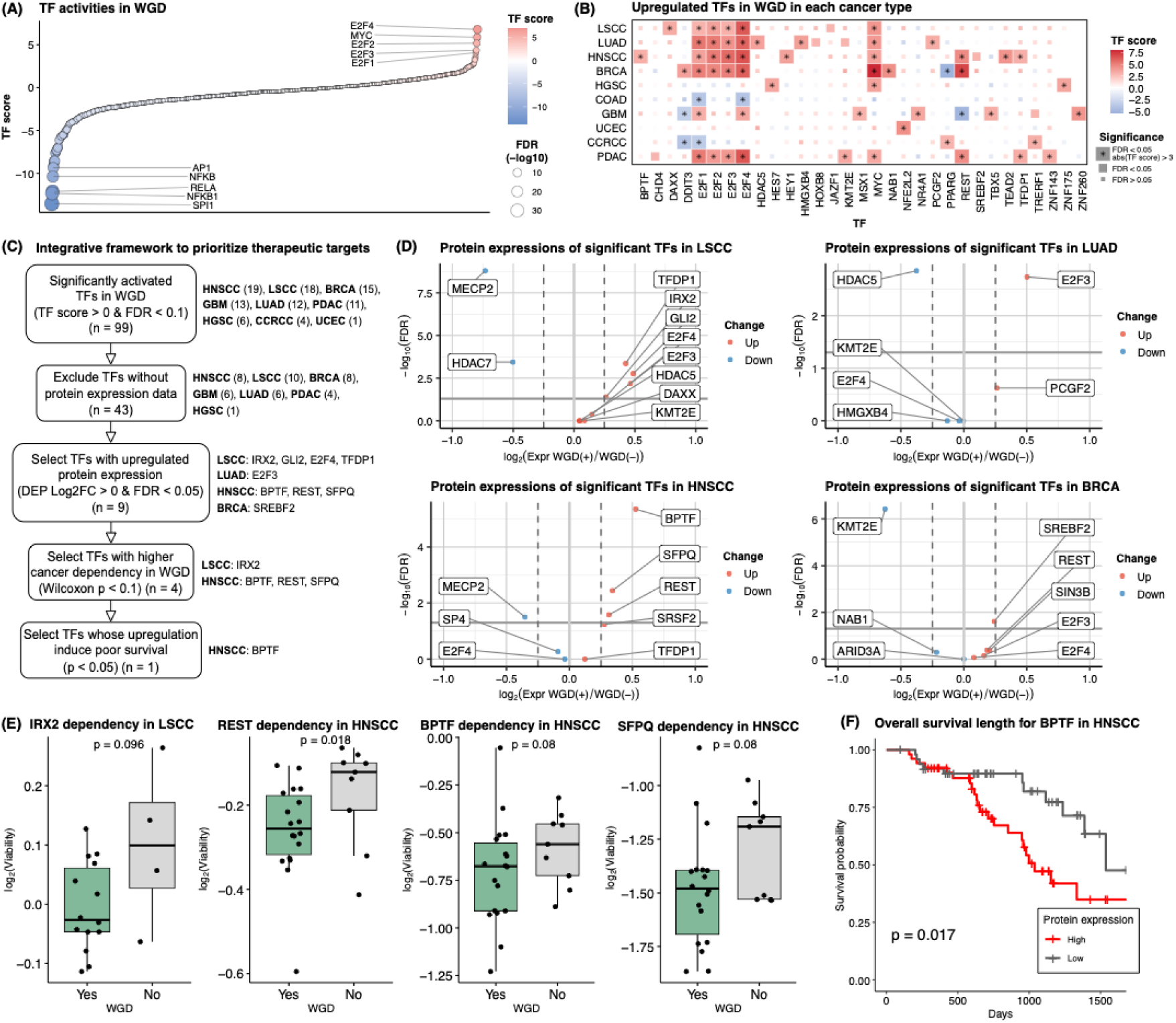
Deciphering potential therapeutic targets among activated TFs in WGD. **(A)** Dot plot depicting estimated TF scores in WGD-positive versus WGD-negative tumors across pan-cancer analysis. The top and bottom five TFs, ranked by TF score, are labeled. **(B)** Significantly upregulated TFs in WGD in each tumor type. The tiles are colored based on the TF estimation score, and TFs meeting the criteria of FDR < 0.05 and TF score > 3 are denoted with asterisks. **(C)** Schematic diagram showing the filtering processes to identify effective therapeutic targets. The number of TFs or the name of TFs remaining after each step is indicated on the right. **(D)** Volcano plots of differentially expressed proteins among significant TFs in WGD-positive tumors compared to WGD-negative tumors. **(E)** Boxplots comparing log_2_ cell viability between WGD-positive and WGD-negative cells after depleting genes with CRISPR in each cancer cell line. **(F)** Kaplan–Meier curve of overall survival based on protein expression levels of BPTF in HNSCC.

Since many TFs have been deemed as ‘undruggable’ due to their structural complexities and a lack of tractable binding sites ^36,37^, we introduced an integrative framework to prioritize WGD-activated TFs as putative therapeutic targets (**Figures 3C and 3SA–I; Table S3C**). Among the 99 TFs that exhibited significant activation in each tumor type (FDR < 0.1), we first selected TFs that were detected in CPTAC proteomics data with high confidence (FDR < 0.01). We then filtered TFs exhibiting significantly upregulated protein expression in WGD-positive tumors compared to WGD-negative tumors. We found that the protein expression of IRX2, GLI2, E2F4, and TFDP1 in LSCC, E2F3 in LUAD, BPTF, REST, and SFPQ in HNSCC, and SREBF2 (also known as SREBP2) in BRCA was significantly upregulated (FDR < 0.05, integrated hypothesis test) (**Figure 3D**). The cancer dependencies of these TFs in each cancer cell line were then evaluated by comparing the dependencies between cells with and without WGD. Among the nine TFs that showed significant upregulation at the protein level, four TFs including IRX2 in LSCC and BPTF, REST, and SFPQ in HNSCC exhibited significantly decreased viability in cells with WGD upon CRISPR-mediated depletion (p < 0.1, Wilcoxon rank-sum test) (**Figure 3E**). Finally, we selected the TFs with high protein expression levels that were associated with poor prognosis. We found a significant association between high BPTF protein expression and unfavorable prognosis in HNSCC (p = 0.017, log-rank test) (**Figure 3F**). BPTF is reported as a co-factor of c-MYC leading to c-MYC-driven proliferation and G1 to S progression ^38^, and we also observed significant activation of MYC in HNSCC (FDR = 5.66×10^−3^). Our findings suggest that deactivating BPTF in patients with WGD-positive HNSCC could slow down cancer cell proliferation, which may ultimately benefit patient survival.

Several TFs did not meet the criteria outlined in our integrative framework; however, they remain potential candidates for further consideration as therapeutic targets. In LSCC, GLI2 showed both significant activation (FDR = 9.37×10^−2^) and upregulation at both mRNA and protein levels (mRNA, FDR = 1.71×10^−8^, Wald test; protein, FDR = 1.72×10^−3^, integrated hypothesis test) (**Figure S3B; Table S3C**). As GLI2 is known to promote cell proliferation and cancer cell survival by upregulating the expression of antiapoptotic proteins in LSCC ^39^, degrading GLI2 may be a promising strategy for the treatment of patients with LSCC harboring WGD. Additionally, TFDP1 and TFDP2, which are partner proteins of the E2F family crucial for the G1 to S phase transition ^40,41^, were identified as potential therapeutic targets for WGD-positive samples in LSCC, HNSCC, PDAC, and HGSC, in which these TFs were significantly activated (FDR < 0.1) (**Figure S3A, S3B, S3E, and S3I**). In LUAD, E2F3 showed significant activation (FDR = 3.60×10^−4^) and upregulation at both mRNA and protein levels (mRNA, FDR = 3.16×10^−5^, Wald test; protein, FDR = 1.83×10^−3^, integrated hypothesis test) (**Figure S3C**). Therefore, E2F3 inhibitors such as Edifoligide could be a potential treatment for LUAD in patients with WGD. Furthermore, in BRCA, SREBF2 showed significant activation (FDR = 8.67×10^−2^) along with upregulated protein expression (FDR = 2.42×10^−2^, integrated hypothesis test) (**Figure S3D**). Targeting SREBF2 could be effective for WGD-positive BRCA samples since SREBF2 is known to regulate the synthesis of cholesterol which is crucial for cancer cell viability in breast cancer cells ^42–44^. Taken together, our integrative framework incorporated protein expression, cancer dependency, and association with patient survival to delineate effective therapeutic targets among significantly activated TFs. Based on our analysis, we propose that BPTF could potentially serve as a therapeutic target for WGD-positive samples in HNSCC, thereby improving patient prognosis.

### Drug repurposing strategies to target key kinases in WGD

As kinases are one of the well-known druggable proteins ^37,45^, we next questioned whether we can suggest treatment strategies by deciphering kinase activities in WGD and matching appropriate drugs targeting these kinases. To estimate kinase activities in the WGD in each tumor type, we utilized a kinase-substrate interaction database ^46^ and phosphoprotein expression data of substrates (see Methods). Consistently, we observed that cancer types with upregulated cell cycle pathways exhibited CDK1 and CDK2 activation (FDR < 0.1) (**Figure 4A; Table S4A**). The protein expression of these cyclin-dependent kinases was also significantly upregulated (FDR < 0.05, integrated hypothesis test) (**Figure 4B**). Apart from cell cycle regulation, kinases involved in the DNA damage response and hypoxia-induced autophagy were significantly activated in some tumors. CSNK2A1, which phosphorylates key components of DNA damage and repair pathways ^47^, was significantly activated in LSCC and LUAD (LSCC, FDR = 7.62×10^−5^; LUAD, FDR = 1.05×10^−3^) (**Figure 4A; Table S4A**). Additionally, in LSCC, PAK4 showed significant activation in WGD (FDR = 6.70×10^−3^). It is known to promote proliferation and suppress apoptosis ^48,49^, and its overexpression has been linked to poor prognosis in NSCLC ^50^. In BRCA, both PRKAA2 (also known as AMPK2) and ULK1 were significantly activated (PRKAA2, FDR = 8.34×10^−2^; ULK1, FDR = 3.21×10^−2^). PRKAA2 induces autophagy during glucose starvation by phosphorylating ULK1 ^51,52^. These results imply that although WGD can drive tumorigenesis by accelerating cell proliferation, it can also induce DNA damage and hypoxia, necessitating kinase activation to inhibit apoptosis, as previously noted ^1–3^.

**Figure 4.**
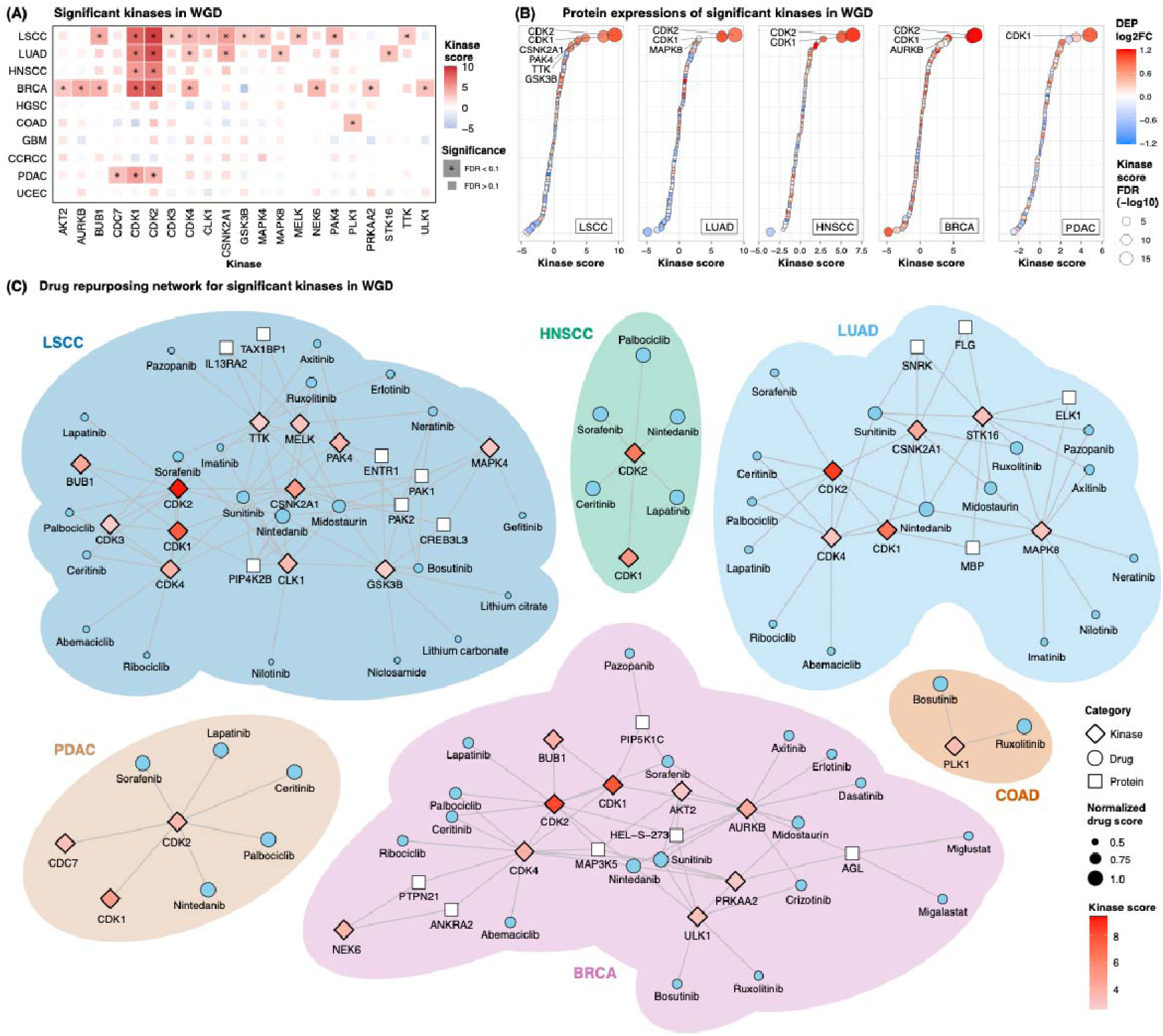
Drug repurposing strategies to target key kinases in WGD. **(A)** Kinases with significant upregulation of expression in association with WGD in each tumor type. The tiles are colored based on the kinase estimation score, and kinases showing significant upregulation are denoted with asterisks. **(B)** Dot plot depicting differentially expressed proteins among significantly upregulated kinases in WGD-positive tumors compared to WGD-negative tumors. Kinases with significant upregulation of protein expression (FDR < 0.05 and log2-fold-change > 0), as well as activation (FDR < 0.1), have been labeled. **(C)** Drug repurposing network for key kinases associated with WGD in each tumor type.

Next, we sought to identify putative drug targets for kinases with upregulated expression in WGD using protein-protein and protein-drug interactions ^53^ (see Methods). Our analysis revealed that nintedanib was the most promising treatment for WGD in LSCC and LUAD, as it interacts with a majority of significantly activated kinases in these cancers (**Figure 4C; Table S4B**). Nintedanib is a small-molecule tyrosine kinase inhibitor that is used for NSCLC patients along with docetaxel after the first line of chemotherapy ^54^. For WGD in COAD, bosutinib and ruxolitinib were suggested to target PLK1 kinase, a pivotal regulator of mitotic events frequently overexpressed in colon cancers ^55–58^. These drugs have been reported to induce apoptosis in colon cancer cells ^59,60^. Sunitinib was recommended as the most suitable drug for BRCA patients with WGD, supported by its efficacy in BRCA reported in some studies ^61,62^, although caution is advised when applying it to metastatic breast cancer patients ^63^. Overall, our findings unraveled cancer-type-specific kinase activations in WGD contributing to cell proliferation, DNA damage response, and hypoxia-driven autophagy. Based on these findings, we proposed FDA-approved drugs for treating patients with WGD in each tumor type.

## Discussion

Our study elucidated the proteogenomic characteristics of WGD in a cancer-type-specific manner. CN signatures were used to characterize WGD tumors into three major groups: WGD type 1, with a highly unstable genome (LSCC, LUAD, and HNSCC); type 2, with focal instability (BRCA and HGSC); and type 3, with tetraploidy (CCRCC, COAD, GBM, PDAC, and UCEC). This classification seems to align with cancer-specific profiles of structural variation ^64^. Triple-negative breast cancer and ovarian cancer are enriched in an SV class, characterized by a high burden of deletions, duplications, and templated insertion chains. LSCC and HNSCC are characterized by an enriched breakage fusion bridge cycle, which is a mechanism of chromosomal instability ^65^. The presence of CN7 and CN15 signatures underscores the occurrence of WGD in a genomically unstable environment at the chromosomal level ^66^.

While the etiology of CN18 remains unknown, we observed its enrichment in WGD-positive BRCA and GBM. CN18 has been linked to TP53 mutation in BRCA ^66^, and our study demonstrates a significant enrichment of TP53 mutation in WGD-positive BRCA samples. This implies CN18 in BRCA may be a WGD signature driven by TP53 mutation. Additionally, CN18 has been associated with hypoxia, which is known to induce polyploid giant cancer cells with stem-like phenotypes in GBM ^67,68^. As a hypoxic condition in GBM has been linked to advanced tumor stage and invasion ^69^, CN18 in GBM may imply hypoxia-driven WGD with an invasive cell state.

In recent studies, WGD tumors have been predominantly characterized by an upregulated cell cycle and a downregulated immune response ^1,2,24,25^. However, our study demonstrated that this pattern is specific to WGD type 1 (LSCC, LUAD, and HNSCC) and does not entirely represent other types of WGD-positive tumors. For these cancer types, significant activation of E2F and MYC seemed to regulate the cell cycle pathway in WGD-positive tumors. Instead, the activities of TFs and kinases were more cancer-type-specific than consistent across WGD types.

Our study has important therapeutic implications for WGD tumors. Given the challenges of targeting undruggable TFs ^36,37,45^, we introduced an integrative approach to identify putative TF targets for new drug development, requiring high protein expression levels, increased cancer dependency, and poorer prognosis in patients with WGD. Based on these criteria, we identified BPTF as a promising target for treating patients with HNSCC harboring WGD. For kinases associated with WGD, we employed a drug repurposing strategy using search algorithms to identify the most suitable drugs targeting these kinases. Drug repositioning is a cost-effective approach for identifying effective treatments without the need for extensive time or resources for new drug development ^70^. We found nintedanib for LSCC and LUAD, bosutinib and ruxolitinib for COAD, and sunitinib for BRCA were recommended as potential drugs for treating WGD patients. Further studies are warranted to evaluate the efficacy of drugs targeting WGD-specific TFs and of repurposed drugs in patient cohorts with WGD. This additional investigation is essential for advancing precision medicine approaches tailored to WGD-associated cancers.

Despite our comprehensive analysis, there are limitations within our study. For cancer types included in CPTAC phase 2 (BRCA, COAD, and HGSC), whole-exome sequencing data were utilized to infer WGD, a method potentially less precise than whole-genome sequencing. In addition, our research covered only 1,060 samples across 10 tumor types, indicating the necessity for larger-scale proteogenomic investigations to fully understand WGD characteristics in various cancers. Future studies should consider employing single-cell and spatial proteomics to gain insights into the cellular dynamics and properties of WGD tumor cells. Furthermore, the effectiveness of BPTF inhibitors in WGD-positive HNSCC, among other drugs for WGD-positive tumors, requires further validation. Despite these limitations, our study on the proteogenomic landscape of WGD across cancers could provide therapeutic insights for treating WGD-related subtypes in individual tumor types, supporting the groundwork for future research endeavors.

## Acknowledgments

This study was supported by the National Research Foundation of Korea (NRF) grant funded by the Korea government (NRF-2019M3E5D3073568 to Joon-Yong An) and by the Ministry of Science and ICT (NRF-2022R1A2C2013377 to Min-Sik Kim), the Korea Health Industry Development Institute (KHIDI) funded by the Ministry of Health & Welfare, Republic of Korea (RS-2023-00304839 to Kwoneel Kim), and Korea University to Joon-Yong An. Eunhyong Chang received a scholarship from the Kwanjeong Educational Foundation and the Brain Korea (BK21) FOUR education program.

## Data availability statement

Public CPTAC datasets are available within 11 publications ^9–18^. Genome and transcriptome data were downloaded from the GDC data portal (https://portal.gdc.cancer.gov). Global proteomic and phosphoproteomic data were downloaded from LinkedOmics (https://www.linkedomics.org/). All data generated during this study are included in Supporting Information.

## Authors’ contributions

Eunhyong Chang and Joon-Yong An designed the study; Eunhyong Chang analyzed the data; Eunhyong Chang, Hee Sang Hwang, Kyu Jin Song, Kwoneel Kim, Min-Sik Kim, Se Jin Jang, Kwang Pyo Kim, Sungyong You, Joon-Yong An wrote and reviewed the manuscript; all authors approved the final version of the manuscript.

## List of abbreviations

WGD: Whole-genome doubling
CIN: Chromosomal instability
BRCA: Breast cancer
CCRCC: Clear cell renal cell carcinoma
COAD: Colon adenocarcinoma
GBM: Glioblastoma
HGSC: High-grade serous carcinoma
HNSCC: Head and neck squamous cell carcinoma
LUAD: Lung adenocarcinoma
LSCC: Lung squamous cell carcinoma
PDAC: Pancreatic ductal adenocarcinoma
UCEC: Uterine corpus endometrial carcinoma
LOH: Loss-of-heterozygosity
TMB: Tumor mutational burden
CN: Copy number
TF: Transcription factor

## Methods

### Data collection and preprocessing

We obtained genomic, transcriptomic, global proteomic, and phosphoproteomic data from the Clinical Proteomic Tumor Analysis Consortium. The mutation annotation format file for each sample and the segment-level copy number variant (CNV) text file for the samples in CPTAC phase-3 (comprising clear cell renal cell carcinoma [CCRCC], GBM, HNSCC, lung squamous cell carcinoma [LSCC], lung adenocarcinoma [LUAD], pancreatic ductal adenocarcinoma [PDAC], and uterine corpus endometrial carcinoma [UCEC]) were downloaded from the GDC data portal (https://portal.gdc.cancer.gov). Transcriptomic data were downloaded from the GDC data portal. Global proteomic and phosphoproteomic data were downloaded from LinkedOmics (https://www.linkedomics.org/).

To standardize the global phosphoprotein data across various cancer types, we initially normalized the data using z-scores. Subsequently, global proteins and phosphoproteins with > 30% missing values in each cancer-type sample were excluded, followed by k-nearest neighbor imputation with k=5 to address missing values.

### CNV calling and WGD detection

As segment-level CN data for BRCA, COAD, and HGSC were unavailable, we acquired BAM files from the GDC data portal and performed the CNV calling process. FACETS v0.16.0 ^71^ was employed to discern allele-specific CN information. The input for FACETS consisted of paired tumor-normal BAM files and a VCF file containing common and germline polymorphic sites downloaded from https://www.ncbi.nlm.nih.gov/variation/docs/human_variation_vcf/.

Using major CN and minor CN data derived from segment-level CNV data, we defined samples as “WGD-positive” if over 50% of their autosomal genome exhibited a major CN greater than or equal to two ^1^.

### Mutational signature analysis

We conducted a mutational signature analysis using the COSMIC signature database v3 ^72^ and R package Sigminer v2.1.5 ^73^. Non-negative matrix factorization was employed to determine the number of signature groups or factorization ranks. This involved creating a tumor-by-component matrix with 50 runs and checking the ranks ranging from 2 to 12. Each signature was identified using the COSMIC signature with the highest cosine similarity. Subsequently, hierarchical clustering was performed, and the samples were assigned to one of the signatures based on the consensus matrix.

### Analysis of differential expression

To discern the differentially expressed genes between WGD-positive and WGD-negative samples, we employed the “DESeq” function from the DESeq2 R package ^74^. The raw count data were used for the generation of a DESeqDataSet using the “DESeqDataSetFromMatrix” function. Following a variance stabilizing transformation and the exclusion of data points with a mean raw count less than 50, we conducted the differential expression analysis utilizing the “DESeq” function, which is grounded in the negative binomial distribution. The DEG analysis was performed using the following formula:

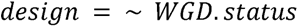

To detect the differential expression of global and phospho-proteins in the WGD samples, we employed an integrated hypothesis testing method following the methodology proposed by Hwang et al ^75^. Briefly, we performed t-tests, median difference tests, and Wilcoxon tests and combined the p-values from the statistical tests using Stouffer’s method. This approach enhances the accuracy of identifying true-positive data elements compared with traditional methods when applied to biological data.

### Gene set enrichment analysis

We conducted a gene set enrichment analysis to elucidate the activated biological pathways in each sample. The “zscore” function from the R package GSVA ^76^ was utilized for analyzing transcriptome, global proteome, and phospho-proteome data. Pathways from various databases, including hallmark (“h.all.v2023.1.Hs.symbols.gmt.txt”), KEGG (“c2.cp.kegg.v2023.1.Hs.symbols.gmt.txt”), Reactome (“c2.cp.reactome.v2023.1.Hs.symbols.gmt.txt”), GO (“c5.go.v2023.1.Hs.symbols.gmt.txt”), and Wikipathway (“c2.cp.wikipathways.v2023.1.Hs.symbols.gmt.txt”) sourced from MSigDB were incorporated into the gene set variation analysis. Subsequently, we performed a t-test between WGD-positive tumors and WGD-negative tumors to identify significantly regulated pathways in the context of WGD.

### Estimation of transcription factor and kinase activity

We used the data corresponding to transcription factor (TF)-target interactions from CollecTRI ^77^ to estimate TF activity. The log-2-fold change values derived from the analysis of differentially expressed genes (DEGs) served as inputs for the identification of TF activity in WGD-positive samples. For estimating kinase activity, we employed kinase-substrate interactions sourced from OmniPathR ^46^. The log-2-fold change values obtained from the analysis of differentially expressed phosphorylated proteins were used as inputs to identify the kinase activity in WGD-positive samples. Subsequently, we employed the “run_mlm” function within the decoupleR R package ^33^ to infer both TF and kinase activities, with the minimum size of regulons set to 1.

### Cancer dependency and druggability analysis

For the comparison of cancer dependencies between WGD-positive and WGD-negative cells, we downloaded “CRISPR (DepMap Public 23Q2+Score, Chronos),” “RNAi (Achilles+DRIVE+Marcotte, DEMETER2),” and “Aneuploidy” data for each cancer cell line from DepMap (https://depmap.org/portal/). We subset the cell lines with annotated genome doubling statuses. Cell lines with one or more instances of genome doubling were classified as WGD-positive, whereas those without genome doubling were categorized as WGD-negative. We conducted a one-sided Wilcoxon rank-sum test to assess whether the WGD-positive cells exhibited lower viability than the WGD-negative cells. To evaluate the druggability of each TF, we used the DGIdb (v5.0.5) ^78^. TFs lacking interactions with drugs were deemed “undruggable,” those with interactions with FDA-approved drugs were designated as “FDA-approved,” and those interacting with known drugs but not with FDA-approved ones were labeled as “druggable.”

### Survival analysis

The Kaplan–Meier estimation model was used to perform survival analysis. The survival duration of the patients and death events were used as inputs for the analysis. To identify the TFs associated with poor prognosis, we compared survival curves between samples with the top 50% and bottom 50% protein expression for each significantly activated TF in WGD (false discovery rate [FDR] < 0.1).

### Drug repurposing network analysis

DrugSt. One (v1.2.0) ^53^ was used to build the drug repurposing network. Significantly upregulated kinases (FDR < 0.1) in the WGD for each tumor type were used as inputs for the search algorithms. NeDRex (v2.21.0) was used for protein-protein and protein-drug interaction searches. For the protein-protein interaction network, we used a multi-Steiner algorithm, setting the number of trees to five, tolerance to five, and hub penalty to 0.5. For the protein-drug interaction network, we used a harmonic centrality algorithm, setting the hub penalty to 0.5 and the result size to 50 and excluding indirect and non-approved drugs.

## Supplemental tables

Table S1.xlsx: Sample overview and copy number signatures associated with WGD

Table S2.xlsx: Enriched pathways in WGD

Table S3.xlsx: Activated transcription factor in WGD and their druggability

Table S4.xlsx: Activated kinases in WGD and drug repurposing network

## Supplemental figures

**Figure S1.**
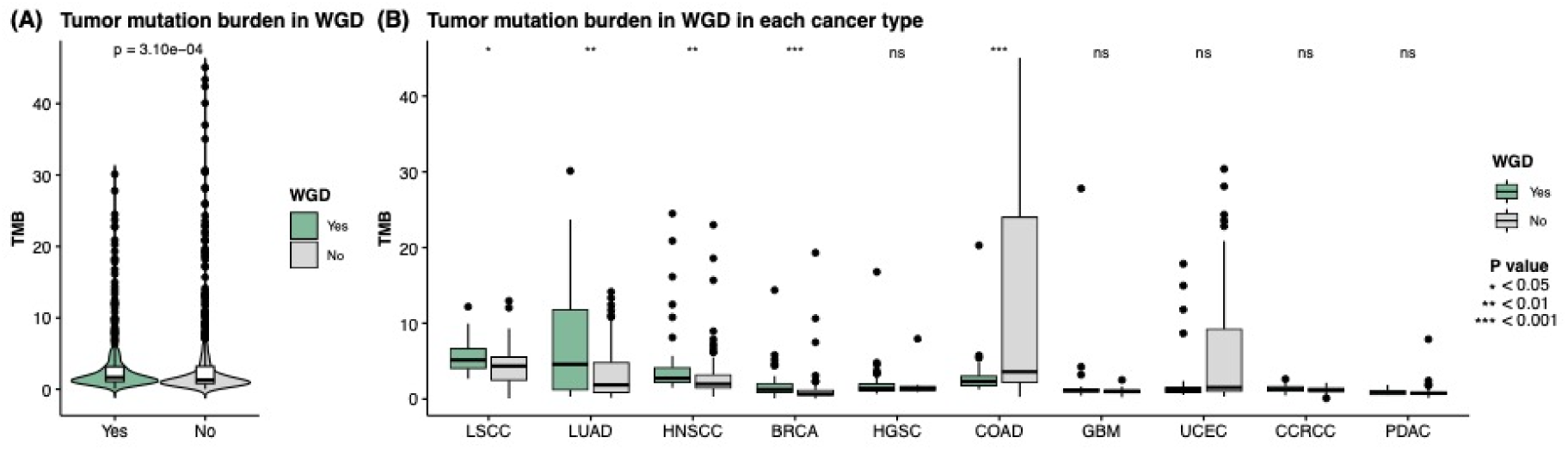
Tumor mutation burden in WGD. **(A)** Violin plot comparing tumor mutation burden between WGD-positive and WGD-negative tumors across 10 cancer types. **(B)** Boxplots comparing tumor mutation burden between WGD-positive and WGD-negative tumors within each cancer type.

**Figure S2.**
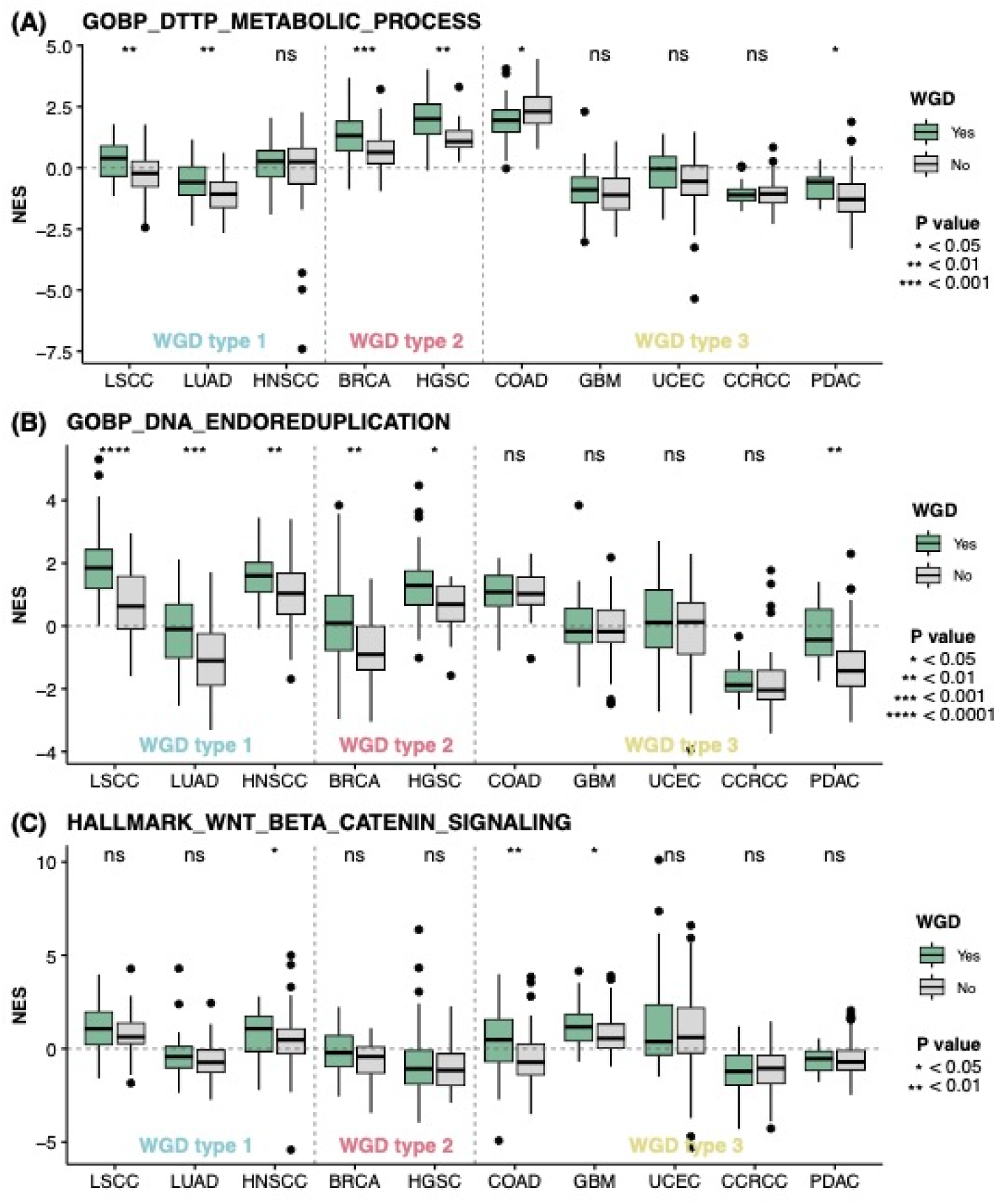
Pathway enrichment in WGD-positive tumors. **(A-C)** Boxplot comparing Normalized enrichment scores (NES) score of dTTP metabolism pathway (GOBP_DTTP_METABOLIC_PROCESS), DNA endoreduplication pathway (GOBP_DNA_ENDOREDUPLICATION), and Wnt signaling pathway (HALLMARK_WNT_BETA_CATENIN_SIGNALING) between WGD-positive and WGD-negative tumors in individual cancer types.

**Figure S3.**
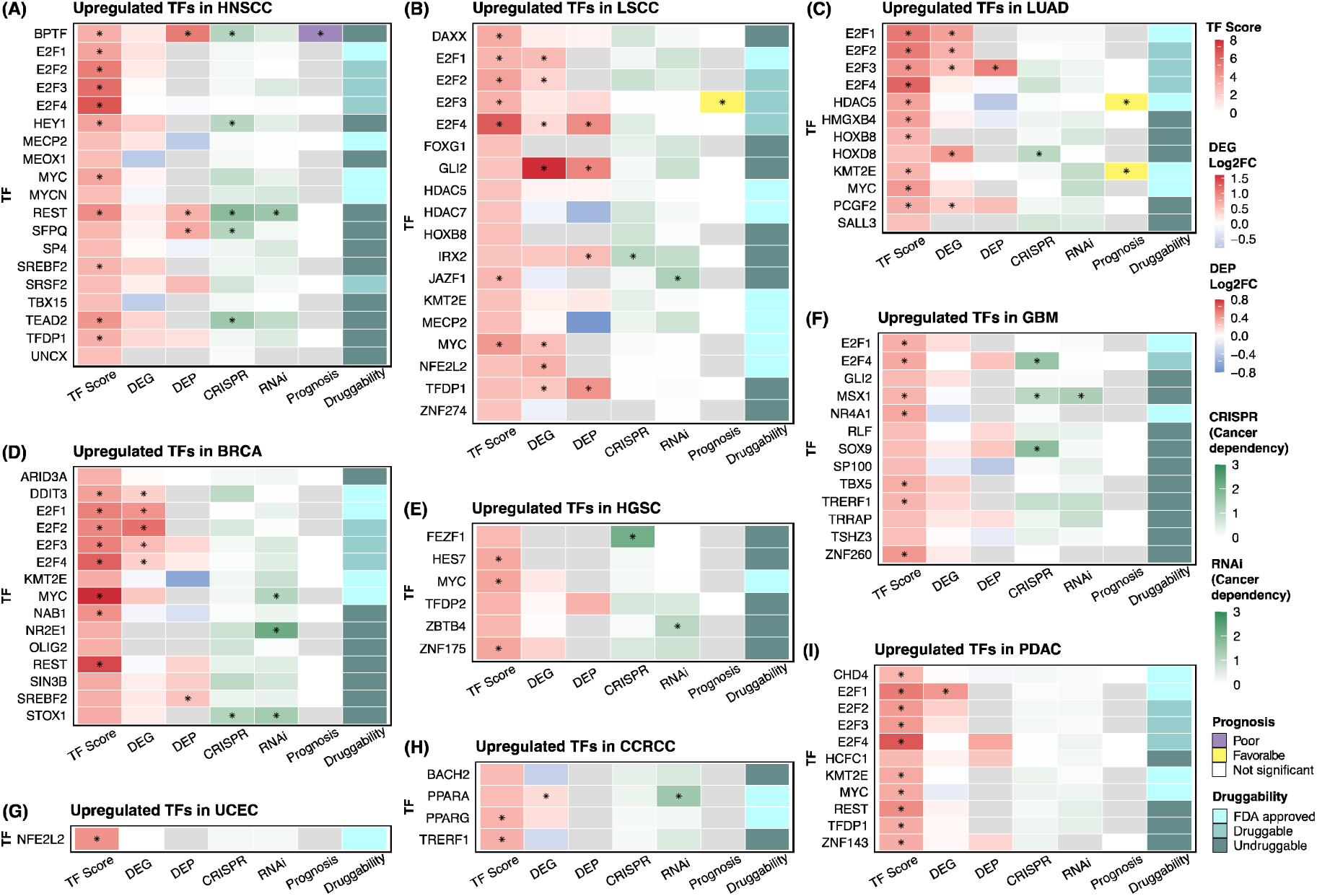
Activated TFs in WGD in each cancer type. **(A-I)** Significantly upregulated TFs in WGD in each cancer type are shown. Significant features are indicated as an asterisk (TF score, FDR < 0.05; DEG, FDR < 0.05; DEP, FDR < 0.05; CRISPR, p < 0.1; RNAi, p < 0.1; Prognosis, p <0.05).

